# The Impact of Face Masks on Performance and Physiological Outcomes during Exercise: A Systematic Review and Meta-analysis

**DOI:** 10.1101/2021.04.22.21255951

**Authors:** Keely A. Shaw, Gordon A. Zello, Scotty J. Butcher, Jong Bum Ko, Leandy Bertrand, Philip D. Chilibeck

**Affiliations:** College of Kinesiology, University of Saskatchewan, Saskatoon, Canada; College of Pharmacy and Nutrition, University of Saskatchewan, Saskatoon, Canada; School of Rehabilitation Science, University of Saskatchewan, Saskatoon, Canada

**Keywords:** Physical Activity, Surgical Mask, COVID-19, N95, Heart Rate

## Abstract

Face masks are promoted for preventing spread of viruses; however, wearing a mask during exercise might increase CO_2_ rebreathing, decrease arterial oxygenation, and decrease exercise performance. A systematic review and meta-analysis was conducted on the impact of wearing a mask during exercise. Data sources included SPORTDiscus, PubMed, and Medline. Eligibility criteria included all study designs comparing surgical, N95, or cloth masks to a no mask condition during any type of exercise where exercise performance and/or physiological parameters were evaluated. Healthy and clinical participants were included. Mean differences (MD) or standardized mean differences (SMD) with 95% confidence intervals were calculated and pooled effects assessed. Twenty-two studies involving 1,573 participants (620 females, 953 males) were included. Surgical, or N95 masks did not impact exercise performance (SMD −0.05 [-0.16,0.07] and −0.16 [-0.54,0.22], respectively) but increased ratings of perceived exertion (RPE) (SMD 0.33 [0.09,0.58] and 0.61 [0.23,0.99]) and dyspnea (SMD 0.6 [0.3,0.9] for all masks). End-tidal CO_2_ (MD 3.3 [1.0, 5.6] and 3.7 [3.0,4.4] mmHg), and heart rate (MD 2 [0,4] beats/min with N95 masks) slightly increased. Face masks can be worn during exercise with no influences on performance and minimal impacts on physiological variables.

**PROSPERO Registration:** CRD42020224988

**Novelty Points:** Face masks can be worn during exercise with no impacts on performance and minimal impacts on physiological variables.

## Introduction

Face masks have been recommended for preventing the spread of viruses (Chu et al., 2020; Hendrix et al. 2020). The potential for spread of infections may be exacerbated during exercise due to heavy breathing, especially indoors such as in fitness (Jang et al., 2020; Lendacki et al., 2021) and sport centres (Atrubin et al., 2020). However, controversy exists when using masks during exercise. While some propose the use of a mask may increase carbon dioxide rebreathing, leading to hypercapnic hypoxia and subsequently decreased tissue oxygenation (Chandrasekaran & Fernandes, 2020), others suggest no impact on exercise (Shaw et al., 2020).

Use of masks during exercise to prevent viral transmission are of importance in 2020-21 due to the COVID-19 pandemic leading to various levels of “lockdown” to prevent virus spread. Measures taken include closure of non-essential services such as fitness and recreation facilities (Douglas et al., 2020); having potential of lasting ill effects on physical and mental health due to increased sedentary behaviour and decreased social contact (Bertrand et al., 2021). Further, physical inactivity increases the risk of developing cardiovascular disease and diabetes, conditions that may exacerbate complications should an individual contract COVID-19 (Bloomgarden, 2020; Nishiga et al., 2020). It is therefore imperative to determine whether wearing a mask during exercise has any important physiological consequences that can affect exercise performance so that when indoor facilities are open, the risk of virus transmission can be minimized.

An expert narrative review recently concluded wearing face masks had minimal impact on physiological function during exercise (Hopkins et al., 2021). We conducted a systematic review and meta-analysis on the impact of wearing a mask during exercise on performance and physiological variables. Based on the review by Hopkins et al. (2021) and observations from our own research group (Shaw et al., 2020), we hypothesized wearing a mask during exercise would have minimal effect on performance and physiological measures.

## Methods

Our systematic review was completed as per the Preferred Reporting Items for Systematic Review and Meta-Analysis (PRISMA) statement (Liberati et al., 2009) and was registered with PROSPERO (CRD42020224988) on December 8^th^, 2020. A literature search was conducted using PubMed, SPORTDiscus, and MedLine including all dates from inception up to March 23, 2021. The following keywords and Boolean phrase were used: (facemask OR face mask OR surgical mask OR N95) AND (exercise OR oxygenation OR physical activity OR heart rate OR rating of perceived exertion OR SpO_2_). A restriction was inclusion of only human trials, but no restrictions on language or date.

The following population, intervention, comparator, outcomes, and study types (PICOS) were included: Any population was considered (i.e. healthy or clinical, any age). The intervention was any variety of mask that is commercially available to the public and might be worn in the general population (i.e. surgical masks, N95 masks, cloth masks). Self-contained breathing apparatuses or masks used in industrial applications and firefighting were not included. Studies with a “no-mask” condition as a comparator were considered. Outcome measures were exercise performance, heart rate, arterial oxygen saturation, oxygen extraction or oxygenation at muscle, arterial partial pressure of carbon dioxide (CO_2_), rating of perceived exertion (RPE), respiratory variables (ventilation, tidal volume, breathing frequency, end-tidal CO_2_), stroke volume, cardiac output, blood pressure, blood lactate, and dyspnea. We considered any study design and only published material. Titles, abstracts, and manuscripts were reviewed by two researchers to determine eligibility based on PICOS parameters. Determination of risk of bias was conducted according to the revised Cochrane risk of bias tool (Stern et al., 2019), using additional considerations for cross-over trials (Higgins et al., 2020). This was completed by two researchers, with disagreements resolved by a third reviewer. Sensitivity analyses were performed by excluding studies with high risk of bias from meta-analyses to determine if they affected results. Reference lists of retrieved articles or any review articles were screened for additional articles.

Data extraction included means and standard deviations (SD) for exercise performance and/or physiological variables for masked and unmasked conditions and were recorded for each study. End-of-exercise means and SDs were used for physiological variables. For example, if the exercise test used was a constant-intensity exercise test for a given duration, variables were extracted from the end of the duration, representing the most stressful time of the exercise test. Likewise, means and SD for physiological variables were extracted from the end of exercise for exercise tests involving a progressive increase in intensity until exhaustion.

Meta-analyses were run by using RevMan 5.3 software (Cochrane Community, London, UK), using fixed-effects models. Heterogeneity was evaluated using *χ*^2^ and *I*^2^ tests where heterogeneity was indicated by either *χ*^2^ *p*-value ≤0.1 or *I*^2^ test value >75%. For most outcome variables, we calculated mean differences (MD) and 95% confidence intervals (CI) between mask and no mask conditions. For the outcomes of exercise performance, dyspnea scale, oxygen extraction at the muscle, and RPE either the units of measurement varied across studies or different scales were used; therefore, we calculated standardized mean differences (SMD) and 95% CI. Study specific MDs or SMDs along with 95% CIs and pooled effects were used to generate forest plots. Publication bias was assessed visually using funnel plots. Surgical and cloth masks are likely to be worn by the general public; whereas, N95 masks are worn mainly by medical personnel and very rarely by the public while exercising. We therefore performed separate meta-analyses for studies evaluating surgical masks and studies evaluating N95 masks when there was enough data for pooling of results. Sensitivity analyses were performed for different types of exercise tests (i.e. maximal or submaximal) and different study populations (i.e. only adults, clinically unhealthy or healthy participants).

## Results

Our search strategy yielded 3,564 articles (Supplementary Figure S1). Twenty-two full text articles (Table 1) with 1,573 participants (620 females, 953 males) met our inclusion criteria. Thirteen of the selected studies were randomized crossover trials, seven were non-randomized crossover trials, and two were retrospective analyses (Table 1).

### Participants

The mean age across studies was 35.6±15.2 years. One study involved children aged 7-14 years (Goh et al., 2019), and the rest adults ≥18 years. All but four studies involved apparently healthy participants (Table 1).

### Exercise Protocols

Seven studies utilized a progressive-intensity exercise test to exhaustion on a cycle ergometer or treadmill, eight studies employed steady-state treadmill or ground walking at constant low-moderate intensity for 6-60 minutes, one used a steady-state constant moderate-vigorous intensity test for 30 minutes on a cycle ergometer, one used a steady state, low intensity cycle ergometer test for 30 minutes, one used a cycle ergometer test for 10 minutes at low intensity and three minutes at moderate intensity, one used treadmill walk at low, moderate, and high intensities for 5-minutes each, two used a 6-minute walk test, and one used a strength training regime (Table 1).

### Masks Used

Fourteen studies utilized a disposable surgical mask, twelve used a variety of N95 masks, two used a cloth mask, and one did not provide sufficient information on the mask used for classification (Table 1). Seven studies investigated more than one type of mask (Table 1).

### Risk of Bias

Of the 22 studies, twelve had high risk of bias, nine had some concerns, and one had low risk of bias (Supplementary Table S1). For the randomization process, some studies either had no randomization (Barbeito-Caamaño et al., 2021; Carrizal and Rodríguez, 2020; Kyung et al., 2020; Laird et al., 2002; Shein et al., 2021), or always had the control condition (no mask) as the first condition, followed by randomization of different mask conditions (Goh et al., 2019; Kim et al., 2013; Roberge et al., 2012a; Roberge et al., 2012b); therefore, were rated as high risk of bias. Studies with “some concerns” for the randomization process had no indication of concealment of the allocation sequence until participants were enrolled and assigned to interventions. For carry-over effects, some studies were considered high risk of bias because they performed the no-mask and mask conditions on the same day with only a short time frame between exercise sessions; therefore, fatigue during one condition could have affected exercise during a subsequent condition. Other studies had “some concerns” for period effects because they did not indicate whether an equal number of participants received the mask or no-mask conditions first. Most studies had “some concerns” with selection of reported results as it could not be determined whether the trial was analyzed with a pre-specified plan (i.e. the study was not registered before participant recruitment).

### Exercise Performance

Nine studies involving 1,156 participants (665 males, 491 females) evaluated exercise performance (Figure 1). Two studies used time to exhaustion, and three used peak power during a progressive-intensity cycle ergometer test (Table 1). Two used metabolic equivalents (METs) achieved during a progressive-intensity test on a treadmill and one used distance achieved on a six-minute walk test (Table 1). A single study assessed power output during a resistance training regime (i.e. 4 sets of 10 repetitions of half-squats at 60% of the one-repetition maximum) (Ramos-Campo et al., 2021). All studies evaluated surgical masks and indicated no effect on performance (Figure 1a; p=0.42). All studies evaluated healthy participants except Carrizal and Rodríguez (2020) and Barbeito-Caamaño et al. (2021), who assessed older adults with a variety of conditions (i.e. Chronic Obstructive Pulmonary Disease COPD, coronary artery disease) during stress testing and Ramos-Campo et al. (2021) who evaluated older individuals with sarcopenia during resistance training. Of the nine studies, three were rated as having high risk of bias; when these were excluded, there was still no effect of wearing a mask on performance (p=0.76). Assessing mode of exercise, there was no effect of wearing a mask during the five cycle ergometer studies (p=0.76) or during the three walking studies (p=0.48). One study evaluated strength training (Ramos-Campo et al., 2021) and found no difference for average power output during a set of squat exercises while wearing a mask compared to no mask (p=0.81).

**Figure 1.**
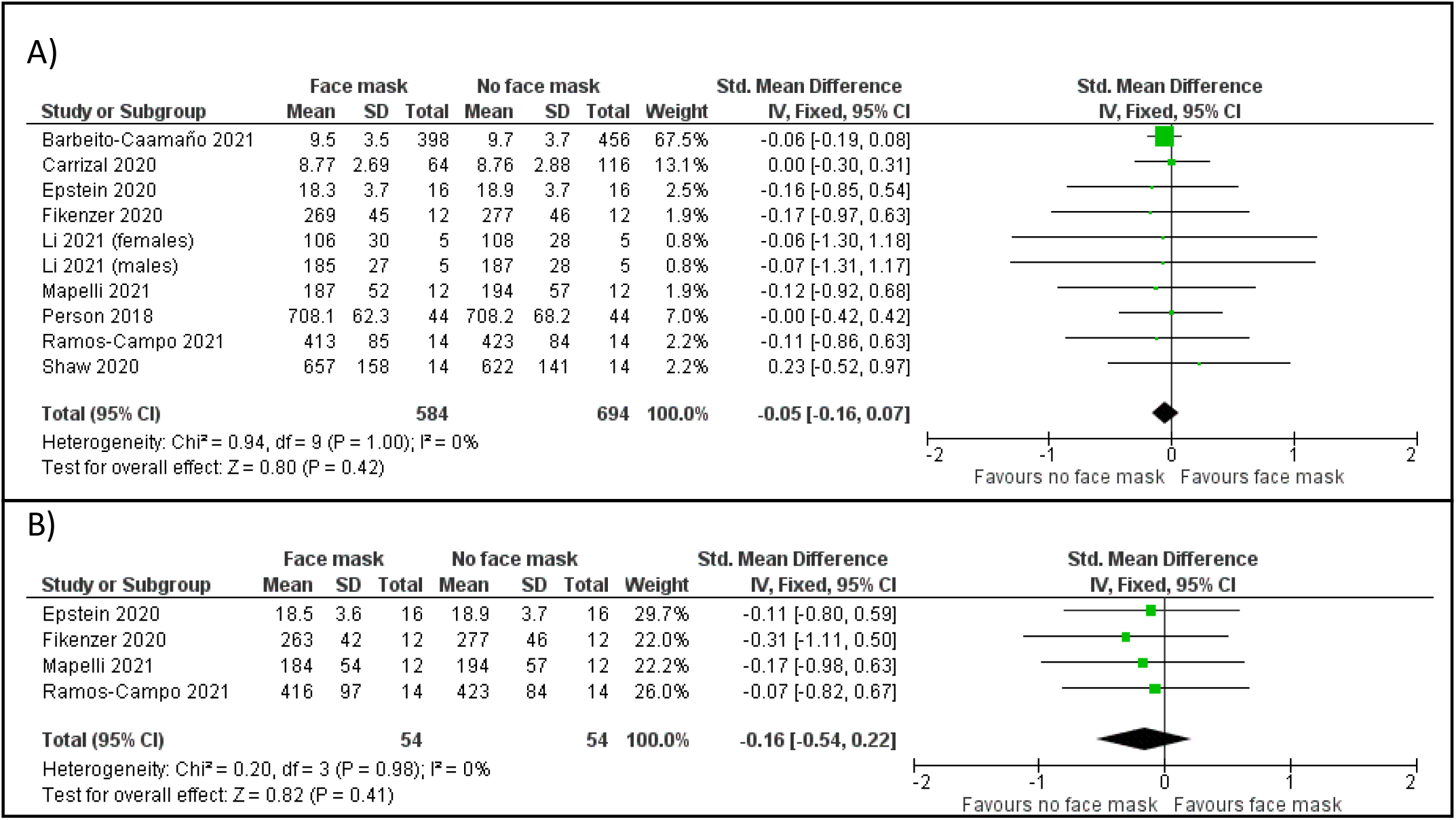
Pooled analysis on the impact of surgical (A) and N95 (B) masks on exercise performance

Four studies also evaluated N95 masks (Table 1) with pooled results indicating no effect on performance (Figure 1b; p=0.41). One study also evaluated cloth masks (Shaw et al., 2020), finding no effect (p=0.49).

### Arterial Oxygen Saturation

Eleven studies involving 386 participants (257 males, 129 females) evaluated impact of a mask on arterial oxygen saturation (Figure 2). Regardless of the mask used, no differences were observed for arterial oxygen saturation during exercise (Figure 2 A-C; surgical mask p=0.09; N95 p=0.07; cloth mask p=0.07). When studies with high risk of bias were excluded from the analyses, there was a small significant reduction in oxygen saturation with N95 (MD = −0.3%; 95% CI −0.6, −0.0%; p=0.03), but not surgical masks (p=0.21). For the studies employing surgical masks, three used submaximal exercise tests (Lässing et al., 2020; Person et al., 2018; Shein et al., 2021) and three used maximal exercise tests (Epstein et al., 2020; Mapelli et al., 2021; Shaw et al., 2020). There was no effect of wearing a surgical mask when only submaximal (p=0.52) tests were included, but a small significant reduction when only maximal tests were included (MD = −0.6%; 95% CI −1.1, −0.0%; p=0.04). For studies using N95 masks, two studies involved maximal exercise testing (Epstein et al., 2020; Mapelli et al., 2021). When only including these, there was a small significant reduction in oxygen saturation (MD = −0.4%; 95% CI −0.7, −0.0%; p=0.03). When only submaximal tests were included, there was no difference (p=0.43). One study using N95 masks involved children (Goh et al., 2018). When this was removed, there was a small, but statistically significant effect of wearing N95 masks (MD = −0.3%; 95% CI −0.6,-0.1%; p=0.006). One study evaluated patients with COPD (Kyung et al., 2020). When removed from the analysis on adults an effect of N95 masks was still present (p=0.03); this result remained significant when studies with high risk of bias were excluded (p=0.03).

**Figure 2.**
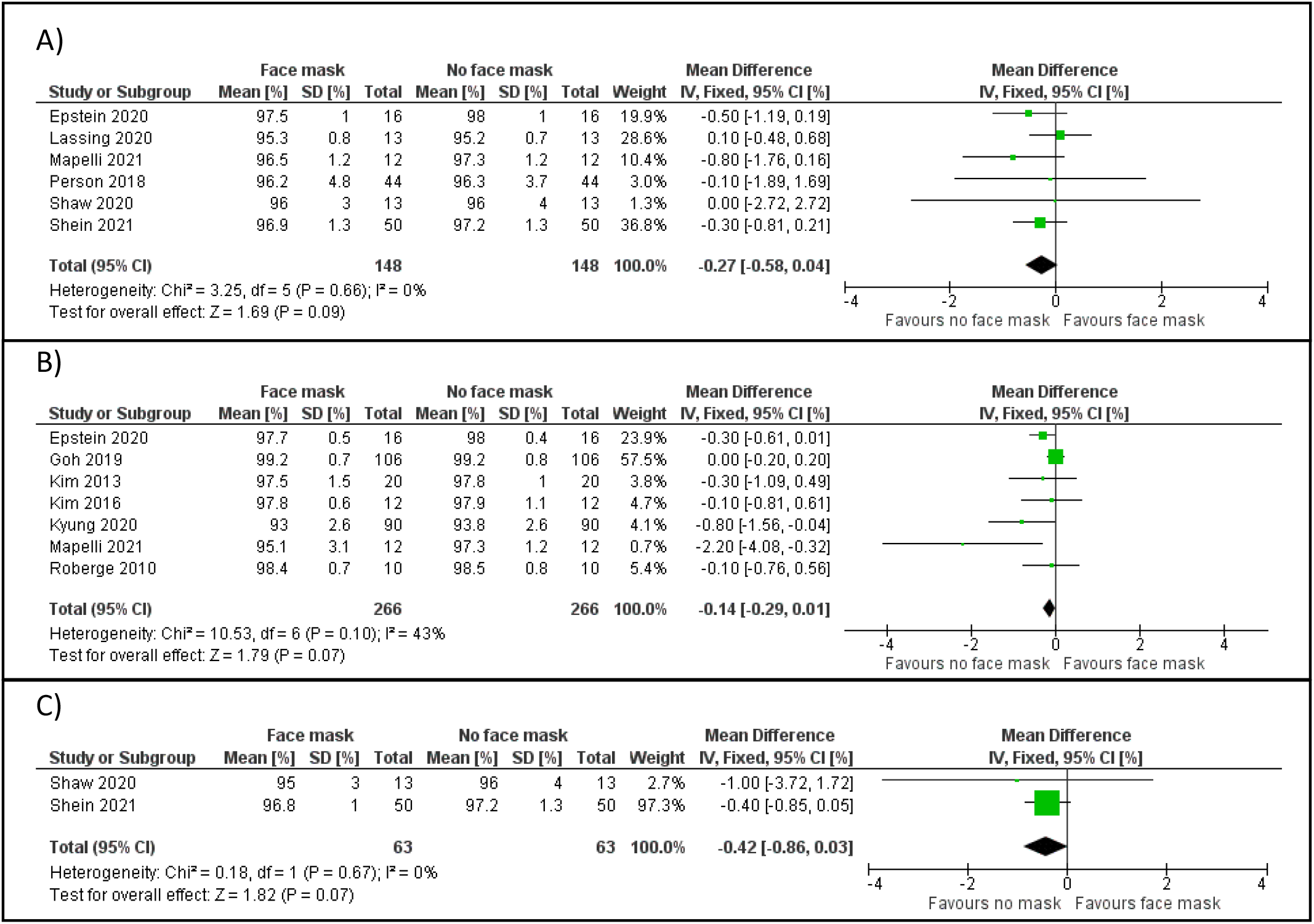
Pooled analysis on the impact of surgical (A), N95 (B), and cloth (C) masks on arterial oxygen saturation.

### Muscle Oxygenation

Three studies involving 40 participants (33 males, 7 females) assessed muscle oxygenation (i.e. muscle oxygen uptake) during exercise while wearing a surgical mask (Supplementary Figure S2). Two evaluated arterial-venous oxygen content difference (Fikenzer et al., 2020; Lässing et al., 2020) and one evaluated tissue oxygenation index (oxygenated hemoglobin relative to total hemoglobin) (Shaw et al., 2020). These studies showed surgical masks had no impact on muscle oxygenation during exercise (p=0.08). Two tests were maximal exercise tests (Fikenzer et al., 2020; Shaw et al., 2020). When only these were included, there was no effect of wearing a surgical mask (p=0.51). Fickenzer et al. (2020) also evaluated N95 masks and found reduced muscle oxygen uptake (p=0.007). Shaw et al. (2020) also assessed a 3-ply cloth mask and found no effect on tissue oxygen index compared to no mask.

### End-tidal and Arterial CO_2_

Arterial CO_2_ was assessed in six studies involving 124 participants (69 males, 55 females) (Supplementary Figure S3; Table 1). For surgical mask studies, Roberge et al. (2012b) did not present SDs, so was excluded from the meta-analysis. Results from the remaining two studies indicated no effect on arterial CO_2_ (p=0.64). Pooled results suggest that exercise while wearing N95 masks leads to significant increases in arterial CO_2_ (Supplementary Figure S3; p=0.05); however, when studies with high risk of bias were excluded, this was no longer significant (p=0.54). Of the studies using N95 masks, only one used maximal exercise testing (Fikenzer et al., 2020). When this study was removed to include only sub-maximal exercise tests, there was no longer an effect of N95 masks (p=00.51). A single study (Shein et al., 2021) assessed the effects of a cloth mask on arterial CO_2_ and found no differences compared to the no mask condition (p=0.16).

End-tidal CO_2_ was assessed in four studies involving 224 participants (166 males, 58 females) (Figure 3 A&B). End-tidal CO_2_ significantly increased when wearing a surgical (p=0.004) or N95 (p<0.001) mask during exercise. Two of these studies with N95 masks had high risk of bias. When removed from analysis, results were still significant (P<0.001). Studies with N95 masks involved a variety of populations and exercise protocols; Epstein et al. (2020) and Mapelli et al. (2021) used a maximal exercise test in healthy adults, Goh et al. (2019) involved children, and Kyung et al. (2020) involved patients with COPD. The results of the meta-analysis were unchanged when each study was removed or if only maximal exercise tests were assessed (p<0.001).

**Figure 3.**
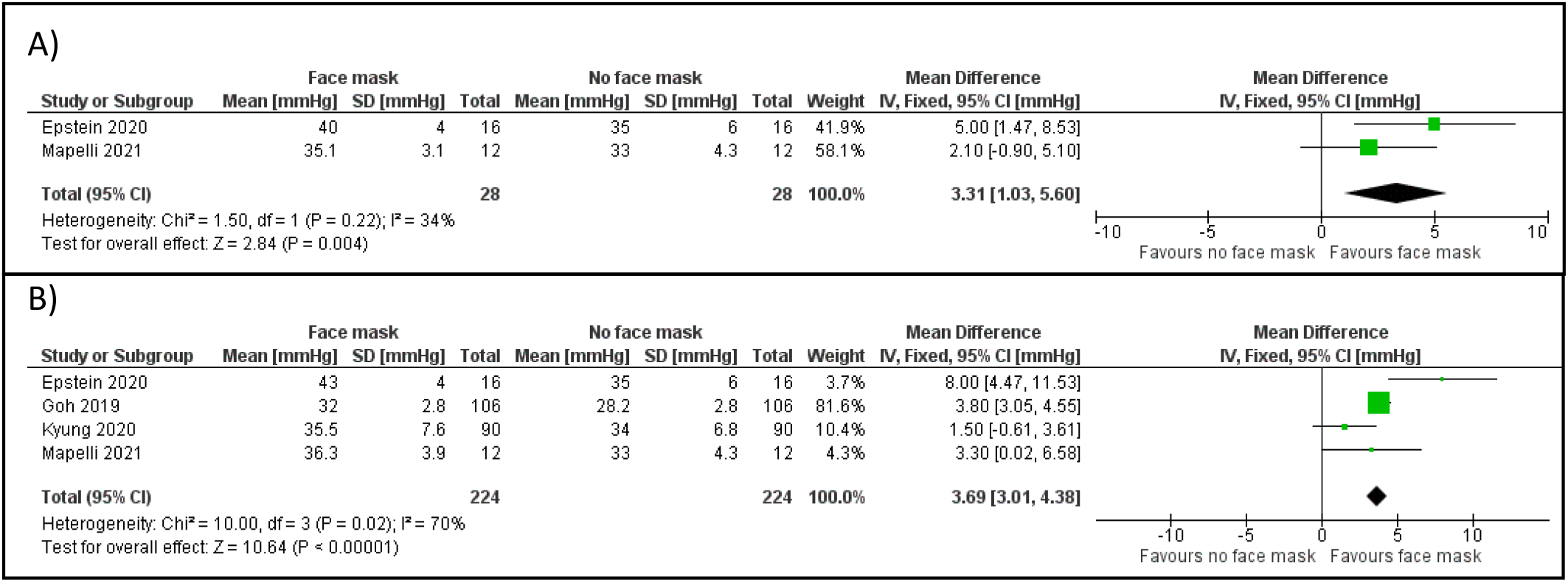
Pooled analysis on the impact of surgical (A) and N95 (B) masks on end-tidal CO_2_

### Ratings of Perceived Exertion

RPE was assessed in 11 studies with 169 participants (67 females, 102 males). Both surgical and N95 masks increased RPE compared to the no mask condition (surgical mask p=0.007; N95 p<0.001; Figure 4 A&B). When studies with high risk of bias were excluded RPE for surgical masks was no longer different compared to the no mask condition (p=0.11), but there was still a difference with N95 masks (p=0.007). When only maximal exercise tests were included, results were no longer significant for surgical masks (p=0.23), but still significant for N95 masks (p=0.02). When only submaximal tests were included, RPE was higher when wearing surgical (p=0.007) or N95 (p=0.01) masks compared to wearing no mask

**Figure 4.**
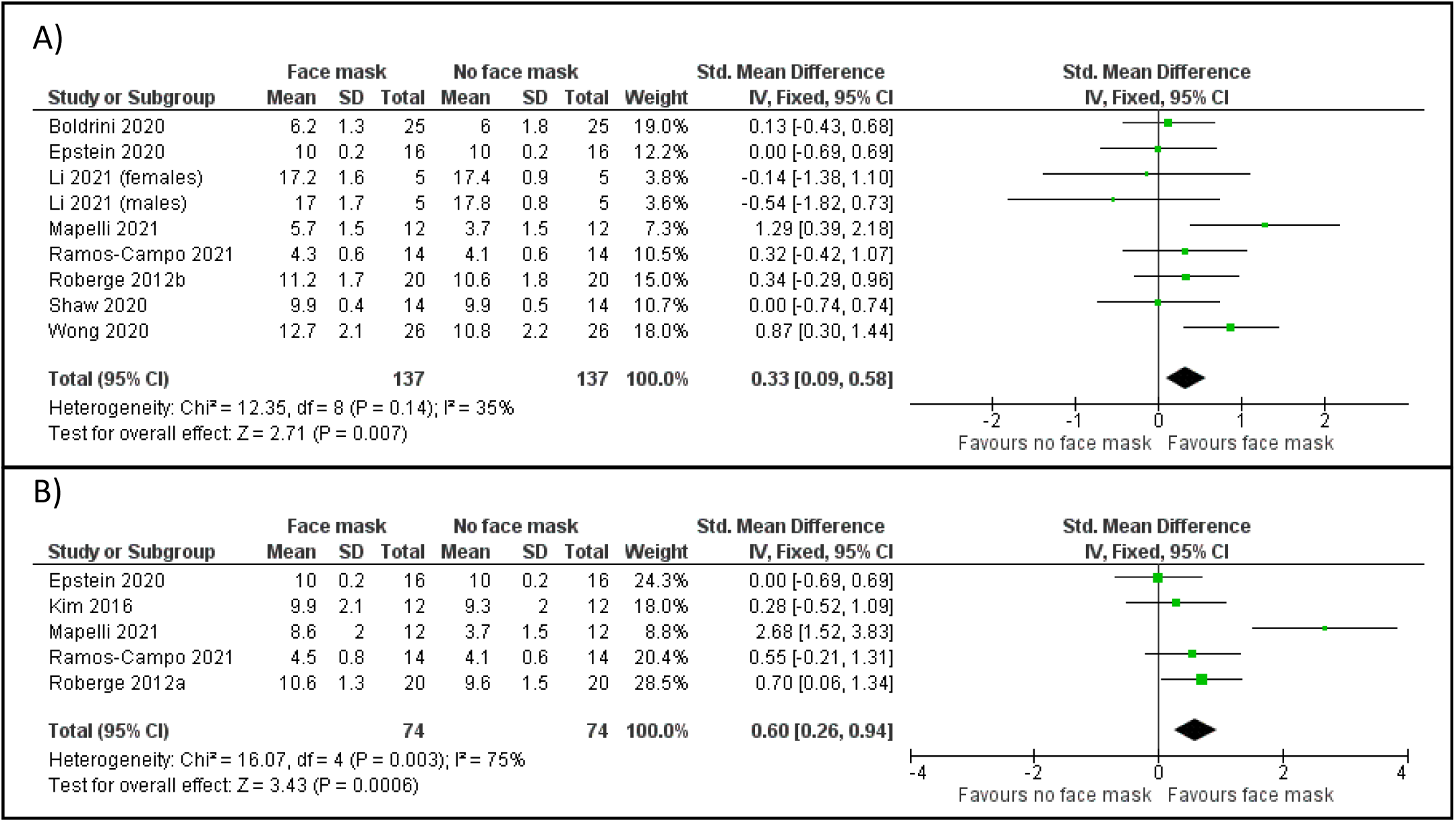
Pooled analysis on the impact of surgical (A) and N95 (B) masks on ratings of perceived exertion

A single study assessed RPE in a strength training setting (Ramos-Campo et al., 2021) and found no difference when wearing either a surgical or N95 mask compared to wearing no mask (p=0.51). A single study assessed wearing a 3-ply cloth mask on RPE during a maximal progressive-intensity cycle ergometer test and found no difference compared to not wearing a mask at any time (Shaw et al., 2020).

### Heart Rate

Twenty studies involving 1,536 participants (925 males, 611 females) (Figure 5; Table 1) evaluated heart rate during exercise while wearing masks. Neither surgical (p=.29) nor cloth (p=.79) masks impacted heart rate (Figure 5 A&C), but the use of N95 masks during exercise increased heart rate compared to not wearing a mask (Figure 5B; p=0.04). When studies with high risk of bias were removed, this was no longer significant (p=0.45,). For studies involving surgical masks, two studies involved older clinical patients, one included sarcopenic adults during strength training, and six involved maximal exercise testing (Table 1). When any combination of different types of studies were removed, heart rate did not differ using surgical masks (p=0.07-0.45). Studies involving N95 masks included three using maximal exercise tests, one involved children, one in patients with COPD, and one that involved strength training in sarcopenic older adults (Table 1). The significant effect of the N95 mask remained when excluding any of these different studies (p=0.03-0.009), but the significant effect was no longer apparent when only studies without high risk of bias were included (p=0.97).

**Figure 5.**
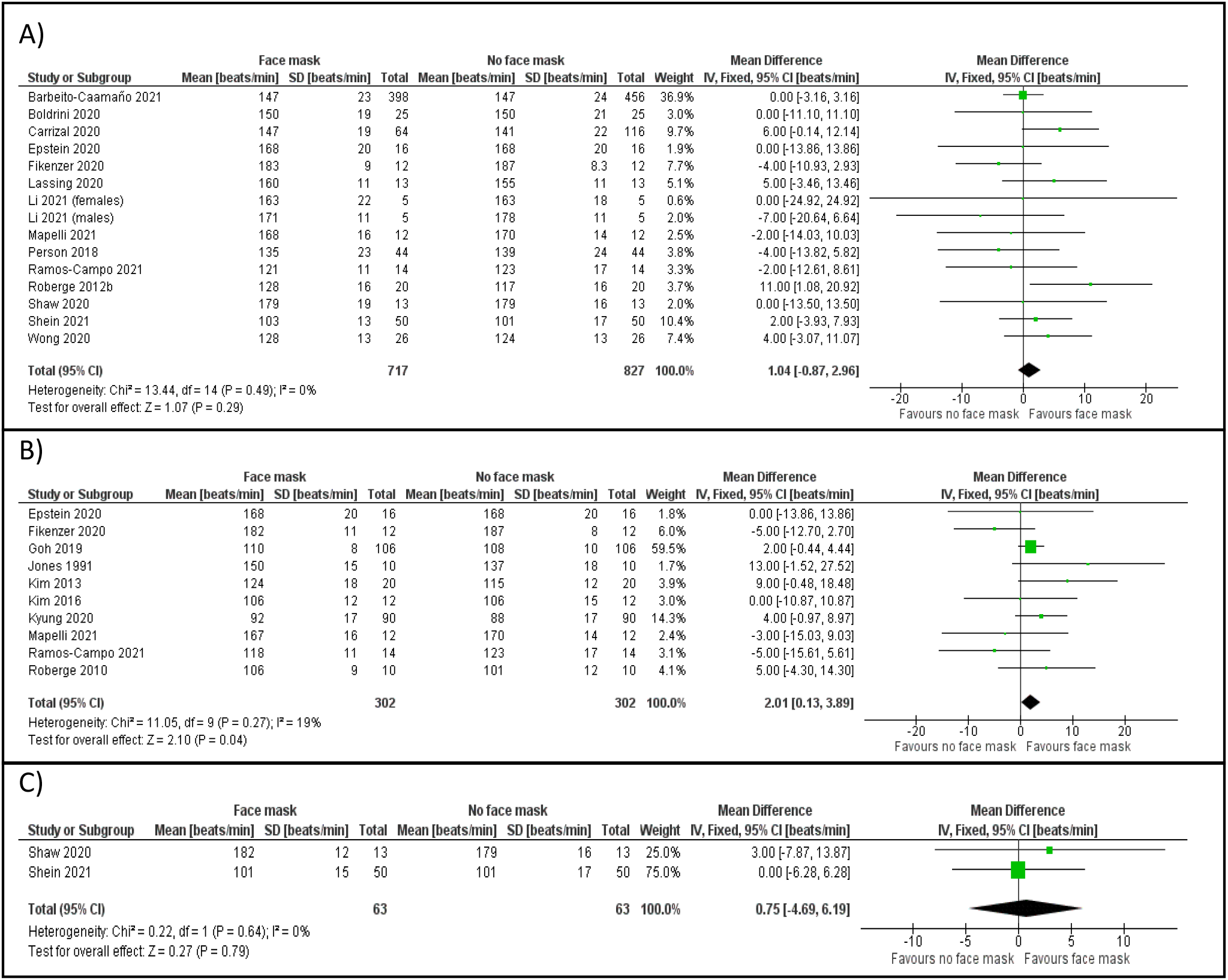
Pooled analysis on the impact of surgical (A), N95 (B), and cloth (C) masks on heart rate

### Cardiac output and Stroke Volume

Two studies involving 26 participants (all males) assessed stroke volume and cardiac output during exercise (supplementary Figures S4 & S5, respectively). Use of surgical masks during exercise had no impact on stroke volume (p=0.14) nor cardiac output (p=0.07). A single study assessing N95 masks found no significant effects on stroke volume (p=0.074) or cardiac output (p=0.42) (Fikenzer et al., 2020).

### Blood Pressure

Seven studies involving 1,178 participants (730 males, 448 females) assessed blood pressure; three of which assessed only systolic blood pressure (supplementary figure S6 A-D). No differences were observed in systolic (p=0.15) or diastolic (p=0.67) blood pressure when a surgical mask was worn during exercise. Similarly, no differences in either systolic (p=0.95) or diastolic (p=0.87) blood pressure were evident when an N95 mask was worn during exercise compared to not wearing a mask. Removal of studies with high risk of bias did not change these results. For studies with a surgical mask, removal of the two studies involving older clinical patients and/or one study involving submaximal exercise testing had no effect on results (p=0.10-0.84). One study using N95 masks involving patients with COPD was the only study using a submaximal exercise test (Kyung et al., 2020). Removing this study did not change results for diastolic (p=0.76) or systolic (p=0.91) blood pressure.

### Dyspnea

Three studies involving 81 participants (55 males, 26 females) assessed dyspnea when wearing either a surgical or N95 mask during exercise (Supplementary Figure S7). Dyspnea was increased when the study using the N95 mask was included (p<0.001) or excluded (p<0.001) from analyses.

### Respiratory Rate

Eleven studies involving 322 participants (243 males, 79 females) assessed respiratory rate (Supplementary Figure S8 A&B). Use of a surgical mask during exercise did not impact respiratory rate (p=0.87) compared to not wearing a mask and was unchanged when the one study with high risk of bias (Roberge et al., 2012b) was excluded (p=0.23). Use of an N95 mask increased respiratory rate (p<0.001), however, significant heterogeneity was present. When Jones et al. (1991) was removed, reducing heterogeneity, the findings were no longer significant whether or not studies with high risk of bias were excluded (p=0.28 and p=0.49, respectively). All sensitivity analyses described below excluded this study. For studies with surgical masks, results did not change when only maximal exercise studies (p=0.33) or submaximal exercise studies (p=0.54) were included. Studies on N95 masks included three that involved maximal exercise testing, one in with COPD, and one in children (Table 1). Exclusion of any combination of these studies resulted in no significant effects (p=0.08-0.51).

### Ventilation and Tidal Volume

Five studies involving 58 participants (40 men, 18 women) assessed ventilation and tidal volume (Supplementary Figures S9 and S10, respectively). Use of a surgical mask during exercise significantly decreased ventilation (p=0.006), but wearing an N95 mask had no impact (p=0.33). When studies with a high risk of bias were removed the result for surgical masks was still significant (p=0.03). Use of neither a surgical (p=0.10) nor N95 (p=0.70) mask had any impact on tidal volume during exercise. This held true when studies with a high risk of bias were removed (p=0.34).

### Lactate

Four studies involving 65 participants (47 males, 18 females) assessed lactate during exercise (Supplementary Figure S11). Surgical masks had no impact on lactate levels during exercise (Supplementary Figure S11A; p=0.48). Of these, one included a maximal cycling test, two involved submaximal cycling, and one included strength training exercise (Table 1). Removal of any combination of these studies still resulted in no impact of masks (p=0.48-0.79). Two studies evaluated lactate levels while wearing N95 masks and found no differences in lactate during exercise (Supplementary Figure S11B; p=0.56).

## Discussion

The most important result of our systematic review and meta-analysis was that wearing a face mask did not significantly affect exercise performance, and that any physiological effects were small. Wearing surgical or N95 masks slightly increased RPE, dyspnea, and end-tidal CO_2_. N95 masks also slightly increased heart rate and respiratory rate. These effects were, however, small and had no impact on exercise performance. For example, although statistically significant, the increase in heart rate while wearing N95 masks for exercise that ranged in intensity from light to maximal was only about 2 beats per minute. Likewise, the increase in respiratory rate while wearing N95 masks during exercise was less than 2 breaths per minute. Although end-tidal CO_2_ was statistically higher while wearing N95 or surgical masks during exercise, values were still within normal physiological ranges across all studies, indicating minimal CO_2_ retention. When studies considered to have high risk of bias were removed from analyses, RPE (for surgical masks) and heart rate (for N95 masks) were no longer different during mask conditions. These findings are in agreement with a recent expert narrative review on the physiological effects of wearing a mask during exercise, which concluded that wearing face masks had little physiological impact (Hopkins et al., 2021).

The use of masks has been recommended by some for prevention of virus spread (Chu et al., 2020; Hendrix et al. 2020); however, the World Health Organization has stated that masks not be worn during vigorous physical activity due to “…the risk of reducing your breathing capacity” (https://www.who.int). This is in contrast to recent recommendations from the Centers for Disease Control and Prevention in the USA who now endorse face masks while exercising in fitness facilities given recent evidence they may reduce virus spread in these facilities (Lendacki et al., 2021). The spread of viruses appears to be heightened in enclosed fitness or sport facilities due to close proximity of individuals, relatively small spaces, and heavy breathing during exercise (Atrubin et al., 2020; Jang et al., 2020; Lendacki et al., 2021).

Our analyses suggest that the use of masks increased RPE, which may be related to increased perceived dyspnea. When studies with high risk of bias were excluded, however, there were no longer differences for RPE when using surgical masks. These studies included multiple testing sessions within a single day with minimal rest between exercise sessions; therefore, fatigue from previous sessions (i.e. carry-over effect) may have increased RPE during mask conditions (Roberge et al., 2012b; Wong et al., 2020).

Our review is strengthened by our assessment of a variety of physiological variables during exercise. In 22 studies analyzed which included 1,573 participants, surgical masks only increased end-tidal CO_2_ and decreased ventilation compared to not wearing a mask, while the use of N95 masks increased arterial and end-tidal CO_2_, heart rate, and respiratory rate. These physiological changes while wearing masks during exercise were small. Some of the physiological changes while wearing masks may also be subject to error. For example, the end-tidal CO_2_ values from Goh et al. (2019) are very low across conditions, therefore their cannula may have been heavily sampling room air. Although end-tidal CO_2_ was significantly higher while wearing N95 or surgical masks from our meta-analysis, all studies were within or below normal ranges, indicating there was most likely minimal CO_2_ retention while wearing a face mask. Many of the physiological measures across studies could be considered “surrogate” measures with methodological shortcomings. For example, measuring arterial oxygen saturation with pulse oximetry is quite variable, end-tidal CO_2_ is a crude estimate of alveolar CO_2_, and most studies estimated arterial CO_2_ levels with transcutaneous measurements, and would not be as accurate as direct measures of blood gases. Ventilation, tidal volume, and respiratory rate may be difficult to measure while wearing a face mask because it necessitates either wearing a mask for breath collection over the face mask (Fikenzer et al., 2020; Lässing et al., 2020; Li et al., 2021; Mapelli et al., 2021) or insertion of a cannula into the nose underneath the face mask (Epstein et al., 2020). This may affect the normal functioning of a face mask during exercise (e.g., wearing a breath-collection mask over a face mask might effectively seal the face mask to the face and not allow air to escape the sides of the mask). Assessment of ventilation using a rubber mask requires that the mask forms an adequate seal with the skin surface of the face to prevent air from escaping. When placing a rubber mask for assessing ventilation over a surgical mask, this seal is lost and air most likely escapes, resulting in lower ventilation measures. A number of studies found marked reductions in ventilation while wearing a surgical mask during maximal cycle ergometry testing but with no significant differences in maximal work rate, supporting this theory (Fikenzer et al., 2020; Li et al., 2021; Mapelli et al., 2021).

Most of the studies we reviewed had either some concerns or high risk of bias from an analysis of trial methodology (Supplementary Table S1); however, others have also criticized the quality of physiological measures and interpretation of some of the articles included in our review (Hopkins et al. 2020). For example, Finkenzer et al. (2020) reported a significant reduction in ventilation, with no change in arterial PCO_2_, a marked decrement in maximal oxygen consumption, but little change of power output and no change in cardiac output. This is not internally consistent, and points to a data collection issue; i.e. likely a leak in their system when testing the face mask conditions.

In conclusion, wearing face masks while exercising has only small effects on physiological responses and no effect on exercise performance. In a long-term pandemic, maintaining a regular exercise routine potentially decreases the risk for conditions such as diabetes and cardiovascular disease, conditions which exacerbate complications due to COVID-19 (Nishiga et al., 2020). Our review found that individuals of various ages and health statuses can wear a mask during exercise with no impacts on performance.

## Data Availability

Research is a Systematic Review. No original data was collected

*The authors declare no conflicts of interest*.

