## Supplementary Table S1 for "The Impact of Face Masks on Performance and Physiological Outcomes during Exercise: A Systematic Review and Meta-analysis"

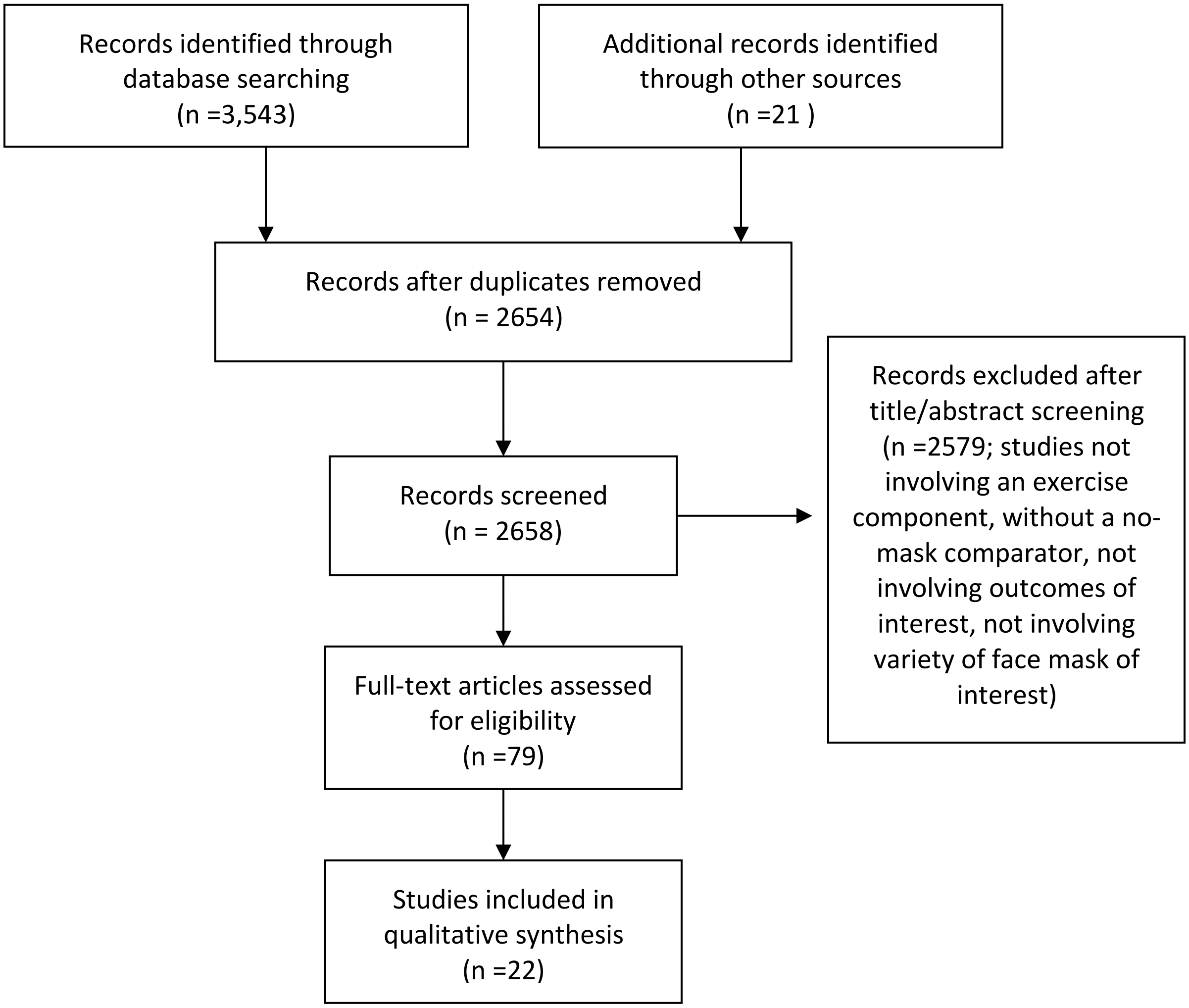


S1. PRISMA diagram: Flow chart of study section process.

| Study or Subgroup | Face mask Mean | Face mask SD | Face mask Total | No face mask Mean | No face mask SD | No face mask Total | Weight | Std. Mean Difference IV, Fixed, 95% CI |
|---|---|---|---|---|---|---|---|---|
| Fikenzer 2020 | 11.5 | 2.2 | 12 | 12.8 | 2.8 | 12 | 31.1% | -0.50 [-1.31, 0.32] |
| Lassing 2020 | 9.6 | 1.4 | 13 | 11.1 | 1.8 | 13 | 31.2% | -0.90 [-1.71, -0.09] |
| Shaw 2020 | -57 | 12 | 14 | -58 | 14 | 14 | 37.6% | 0.07 [-0.67, 0.82] |
| **Total (95% CI)** | | | **39** | | | **39** | **100.0%** | **-0.41 [-0.86, 0.05]** |

Heterogeneity: $Chi^2 = 3.09$, $df = 2$ ($P = 0.21$); $I^2 = 35\%$

Test for overall effect: $Z = 1.76$ ($P = 0.08$)

Std. Mean Difference
IV, Fixed, 95% CI

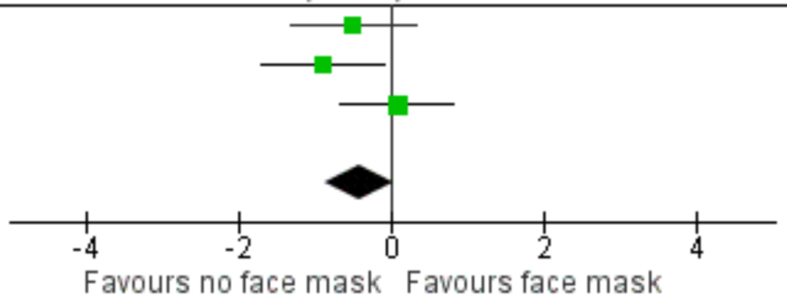


Supplementary Figure S2. Pooled analysis on the impact of masks on tissue oxygenation index

A)

| Study or Subgroup | Face mask Mean [mmHg] | Face mask SD [mmHg] | Face mask Total | No face mask Mean [mmHg] | No face mask SD [mmHg] | No face mask Total | Weight | Mean Difference IV, Fixed, 95% CI [mmHg] |
|---|---|---|---|---|---|---|---|---|
| Fikenzer 2020 | 34 | 6 | 12 | 34 | 4 | 12 | 18.1% | 0.00 [-4.08, 4.08] |
| Shein 2021 | 39.6 | 4.8 | 50 | 40.1 | 5 | 50 | 81.9% | -0.50 [-2.42, 1.42] |
| **Total (95% CI)** | | | **62** | | | **62** | **100.0%** | **-0.41 [-2.15, 1.33]** |

Heterogeneity: Chi² = 0.05, df = 1 (P = 0.83); I² = 0%
Test for overall effect: Z = 0.46 (P = 0.64)

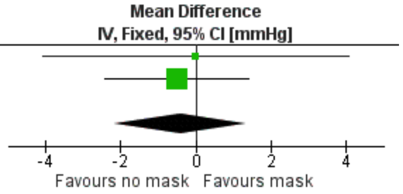


B)

| Study or Subgroup | Face mask Mean [mmHg] | Face mask SD [mmHg] | Face mask Total | No face mask Mean [mmHg] | No face mask SD [mmHg] | No face mask Total | Weight | Mean Difference IV, Fixed, 95% CI [mmHg] |
|---|---|---|---|---|---|---|---|---|
| Fikenzer 2020 | 35 | 5 | 12 | 34 | 4 | 12 | 13.1% | 1.00 [-2.62, 4.62] |
| Kim 2013 | 42.7 | 4 | 20 | 39.7 | 3.8 | 20 | 29.4% | 3.00 [0.58, 5.42] |
| Kim 2016 | 41.3 | 2.4 | 12 | 40.9 | 2.4 | 12 | 46.7% | 0.40 [-1.52, 2.32] |
| Roberge 2010 | 42 | 5.6 | 10 | 40.8 | 3.2 | 10 | 10.8% | 1.20 [-2.80, 5.20] |
| **Total (95% CI)** | | | **54** | | | **54** | **100.0%** | **1.33 [0.02, 2.64]** |

Heterogeneity: Chi² = 2.77, df = 3 (P = 0.43); I² = 0%
Test for overall effect: Z = 1.99 (P = 0.05)

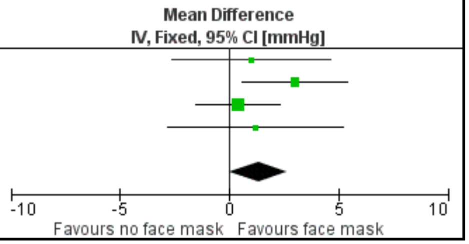


Supplementary Figure S3. Pooled analysis on the impact of surgical (A) and N95 (B) masks on arterial $CO_2$

| | Face mask | | | No face mask | | | | Mean Difference |
|---|---|---|---|---|---|---|---|---|
| **Study or Subgroup** | **Mean [ml]** | **SD [ml]** | **Total** | **Mean [ml]** | **SD [ml]** | **Total** | **Weight** | **IV, Fixed, 95% CI [ml]** |
| Fikenzer 2020 | 165 | 35 | 12 | 151 | 26 | 12 | 44.3% | 14.00 [-10.67, 38.67] |
| Lassing 2020 | 179 | 26 | 13 | 168 | 31 | 13 | 55.7% | 11.00 [-10.99, 32.99] |
| **Total (95% CI)** | | | **25** | | | **25** | **100.0%** | **12.33 [-4.09, 28.75]** |

Heterogeneity: Chi² = 0.03, df = 1 (P = 0.86); I² = 0%
Test for overall effect: Z = 1.47 (P = 0.14)

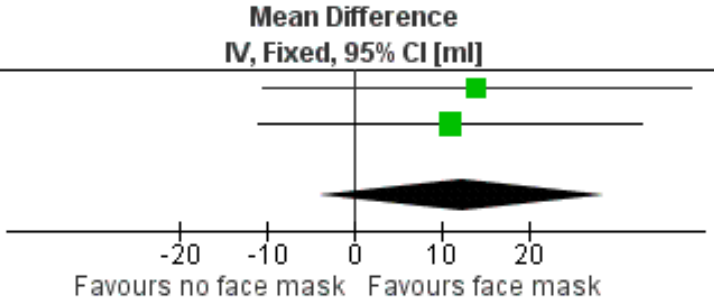


Supplementary Figure S4. Pooled analysis on the impact of masks on stroke volume

| | Face mask | | | No face mask | | | | Mean Difference |
|---|---|---|---|---|---|---|---|---|
| **Study or Subgroup** | **Mean [L/min]** | **SD [L/min]** | **Total** | **Mean [L/min]** | **SD [L/min]** | **Total** | **Weight** | **IV, Fixed, 95% CI [L/min]** |
| Fikenzer 2020 | 27.3 | 5.6 | 12 | 25.8 | 4.2 | 12 | 37.0% | 1.50 [-2.46, 5.46] |
| Lassing 2020 | 28.6 | 3.9 | 13 | 25.9 | 4 | 13 | 63.0% | 2.70 [-0.34, 5.74] |
| **Total (95% CI)** | | | **25** | | | **25** | **100.0%** | **2.26 [-0.15, 4.67]** |

Heterogeneity: Chi² = 0.22, df = 1 (P = 0.64); I² = 0%

Test for overall effect: Z = 1.83 (P = 0.07)

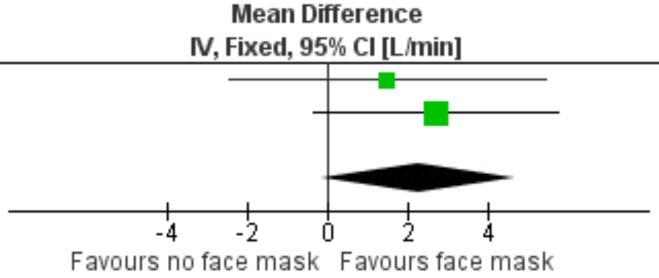


Supplementary Figure S5. Pooled analysis on the impact of masks on cardiac output

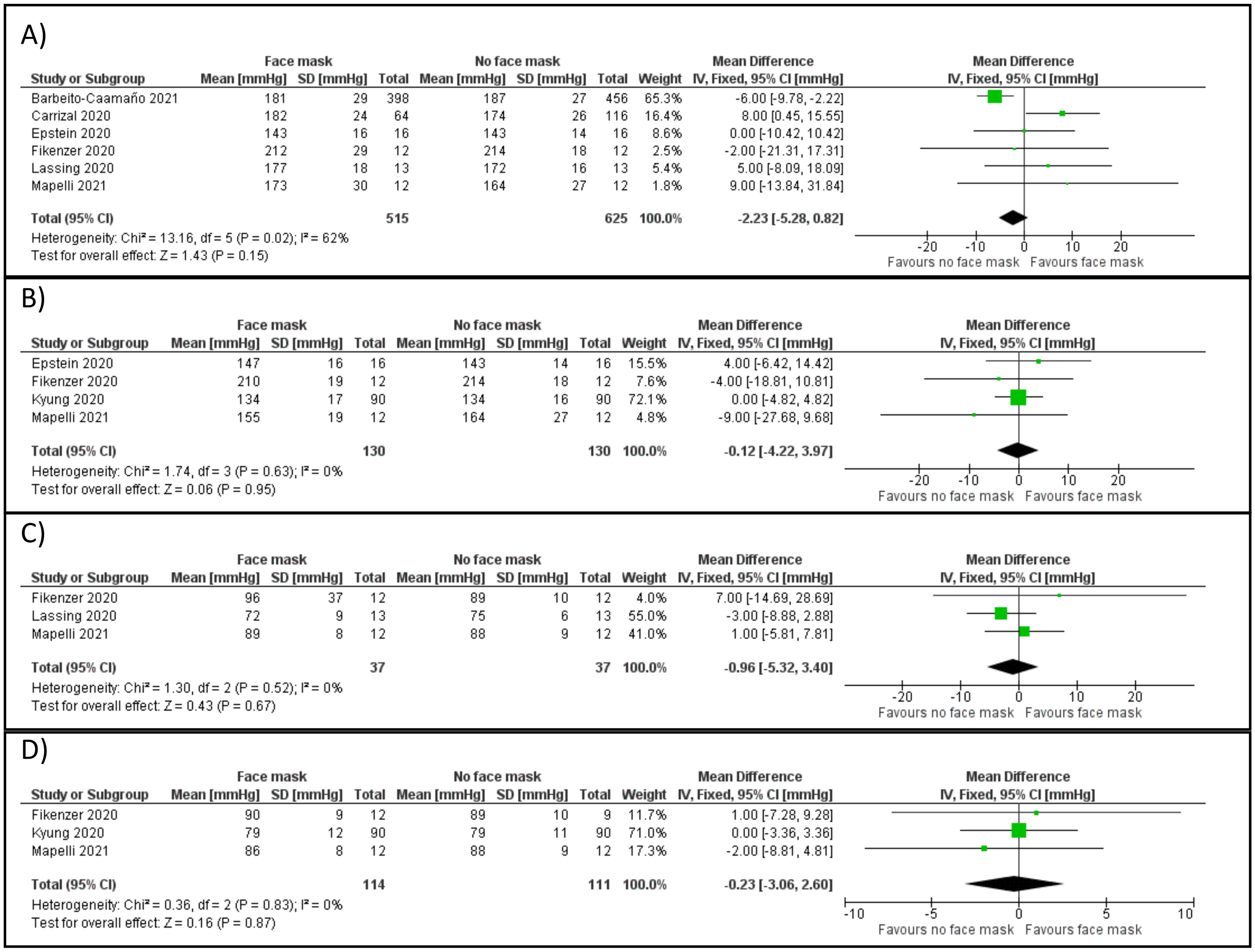


A)

| Study or Subgroup | Face mask Mean [mmHg] | Face mask SD [mmHg] | Face mask Total | No face mask Mean [mmHg] | No face mask SD [mmHg] | No face mask Total | Weight | Mean Difference IV, Fixed, 95% CI [mmHg] |
|---|---|---|---|---|---|---|---|---|
| Barbeito-Caamaño 2021 | 181 | 29 | 398 | 187 | 27 | 456 | 65.3% | -6.00 [-9.78, -2.22] |
| Carrizal 2020 | 182 | 24 | 64 | 174 | 26 | 116 | 16.4% | 8.00 [0.45, 15.55] |
| Epstein 2020 | 143 | 16 | 16 | 143 | 14 | 16 | 8.6% | 0.00 [-10.42, 10.42] |
| Fikenzer 2020 | 212 | 29 | 12 | 214 | 18 | 12 | 2.5% | -2.00 [-21.31, 17.31] |
| Lassing 2020 | 177 | 18 | 13 | 172 | 16 | 13 | 5.4% | 5.00 [-8.09, 18.09] |
| Mapelli 2021 | 173 | 30 | 12 | 164 | 27 | 12 | 1.8% | 9.00 [-13.84, 31.84] |
| **Total (95% CI)** | | | **515** | | | **625** | **100.0%** | **-2.23 [-5.28, 0.82]** |

Heterogeneity: Chi² = 13.16, df = 5 (P = 0.02); I² = 62%
Test for overall effect: Z = 1.43 (P = 0.15)

B)

| Study or Subgroup | Face mask Mean [mmHg] | Face mask SD [mmHg] | Face mask Total | No face mask Mean [mmHg] | No face mask SD [mmHg] | No face mask Total | Weight | Mean Difference IV, Fixed, 95% CI [mmHg] |
|---|---|---|---|---|---|---|---|---|
| Epstein 2020 | 147 | 16 | 16 | 143 | 14 | 16 | 15.5% | 4.00 [-6.42, 14.42] |
| Fikenzer 2020 | 210 | 19 | 12 | 214 | 18 | 12 | 7.6% | -4.00 [-18.81, 10.81] |
| Kyung 2020 | 134 | 17 | 90 | 134 | 16 | 90 | 72.1% | 0.00 [-4.82, 4.82] |
| Mapelli 2021 | 155 | 19 | 12 | 164 | 27 | 12 | 4.8% | -9.00 [-27.68, 9.68] |
| **Total (95% CI)** | | | **130** | | | **130** | **100.0%** | **-0.12 [-4.22, 3.97]** |

Heterogeneity: Chi² = 1.74, df = 3 (P = 0.63); I² = 0%
Test for overall effect: Z = 0.06 (P = 0.95)

C)

| Study or Subgroup | Face mask Mean [mmHg] | Face mask SD [mmHg] | Face mask Total | No face mask Mean [mmHg] | No face mask SD [mmHg] | No face mask Total | Weight | Mean Difference IV, Fixed, 95% CI [mmHg] |
|---|---|---|---|---|---|---|---|---|
| Fikenzer 2020 | 96 | 37 | 12 | 89 | 10 | 12 | 4.0% | 7.00 [-14.69, 28.69] |
| Lassing 2020 | 72 | 9 | 13 | 75 | 6 | 13 | 55.0% | -3.00 [-8.88, 2.88] |
| Mapelli 2021 | 89 | 8 | 12 | 88 | 9 | 12 | 41.0% | 1.00 [-5.81, 7.81] |
| **Total (95% CI)** | | | **37** | | | **37** | **100.0%** | **-0.96 [-5.32, 3.40]** |

Heterogeneity: Chi² = 1.30, df = 2 (P = 0.52); I² = 0%
Test for overall effect: Z = 0.43 (P = 0.67)

D)

| Study or Subgroup | Face mask Mean [mmHg] | Face mask SD [mmHg] | Face mask Total | No face mask Mean [mmHg] | No face mask SD [mmHg] | No face mask Total | Weight | Mean Difference IV, Fixed, 95% CI [mmHg] |
|---|---|---|---|---|---|---|---|---|
| Fikenzer 2020 | 90 | 9 | 12 | 89 | 10 | 9 | 11.7% | 1.00 [-7.28, 9.28] |
| Kyung 2020 | 79 | 12 | 90 | 79 | 11 | 90 | 71.0% | 0.00 [-3.36, 3.36] |
| Mapelli 2021 | 86 | 8 | 12 | 88 | 9 | 12 | 17.3% | -2.00 [-8.81, 4.81] |
| **Total (95% CI)** | | | **114** | | | **111** | **100.0%** | **-0.23 [-3.06, 2.60]** |

Heterogeneity: Chi² = 0.36, df = 2 (P = 0.83); I² = 0%
Test for overall effect: Z = 0.16 (P = 0.87)

Supplementary Figure S6. Pooled analysis on the impact of masks on blood pressure. A= surgical masks on systolic blood pressure; B= N95 masks on diastolic blood pressure; C= surgical masks on systolic blood pressure; D= N95 masks on diastolic blood pressure

| Study or Subgroup | Face mask Mean | Face mask SD | Face mask Total | No face mask Mean | No face mask SD | No face mask Total | Weight | Std. Mean Difference IV, Fixed, 95% CI |
|---|---|---|---|---|---|---|---|---|
| Boldrini 2020 | 5.7 | 2 | 25 | 4.6 | 2.3 | 25 | 31.3% | 0.50 [-0.06, 1.07] |
| Kim 2016 | 2.2 | 0.6 | 12 | 1.7 | 0.8 | 12 | 14.5% | 0.68 [-0.14, 1.51] |
| Person 2018 | 5.6 | 1.8 | 44 | 4.6 | 1.3 | 44 | 54.2% | 0.63 [0.20, 1.06] |
| **Total (95% CI)** | | | **81** | | | **81** | **100.0%** | **0.60 [0.28, 0.91]** |

Heterogeneity: Chi² = 0.17, df = 2 (P = 0.92); I² = 0%
Test for overall effect: Z = 3.72 (P = 0.0002)

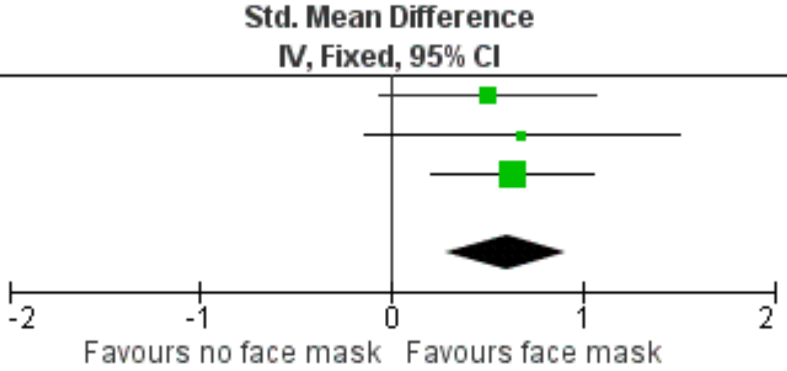


Supplementary Figure S7. Pooled analysis on the impact of masks on dyspnea

A)

| Study or Subgroup | Face mask Mean [breath/min] | Face mask SD [breath/min] | Face mask Total | No face mask Mean [breath/min] | No face mask SD [breath/min] | No face mask Total | Weight | Mean Difference IV, Fixed, 95% CI [breath/min] |
|---|---|---|---|---|---|---|---|---|
| Epstein 2020 | 36.5 | 4 | 16 | 36.5 | 4 | 16 | 25.8% | 0.00 [-2.77, 2.77] |
| Fikenzer 2020 | 39 | 6 | 12 | 41 | 5 | 12 | 10.2% | -2.00 [-6.42, 2.42] |
| Lassing 2020 | 32.1 | 5.4 | 13 | 34 | 7.3 | 13 | 8.1% | -1.90 [-6.84, 3.04] |
| Mapelli 2021 | 37.7 | 5.5 | 12 | 41.5 | 8 | 12 | 6.6% | -3.80 [-9.29, 1.69] |
| Roberge 2012b | 24.7 | 3.7 | 20 | 23.7 | 2.7 | 20 | 49.3% | 1.00 [-1.01, 3.01] |
| **Total (95% CI)** | | | **73** | | | **73** | **100.0%** | **-0.12 [-1.52, 1.29]** |

Heterogeneity: Chi² = 4.12, df = 4 (P = 0.39); I² = 3%

Test for overall effect: Z = 0.16 (P = 0.87)

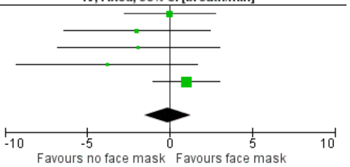


B)

| Study or Subgroup | Face mask Mean [breath/min] | Face mask SD [breath/min] | Face mask Total | No face mask Mean [breath/min] | No face mask SD [breath/min] | No face mask Total | Weight | Mean Difference IV, Fixed, 95% CI [breath/min] |
|---|---|---|---|---|---|---|---|---|
| Epstein 2020 | 37 | 4 | 16 | 36.5 | 4 | 16 | 6.0% | 0.50 [-2.27, 3.27] |
| Fikenzer 2020 | 37 | 6 | 12 | 41 | 5 | 12 | 2.4% | -4.00 [-8.42, 0.42] |
| Goh 2019 | 23.2 | 3.7 | 106 | 23.5 | 3.5 | 106 | 49.1% | -0.30 [-1.27, 0.67] |
| Jones 1991 | 30 | 2 | 10 | 20 | 2 | 10 | 15.0% | 10.00 [8.25, 11.75] |
| Kim 2013 | 23.6 | 5.8 | 20 | 21.7 | 3.4 | 20 | 5.3% | 1.90 [-1.05, 4.85] |
| Kim 2016 | 28.4 | 3.2 | 12 | 28.1 | 7.1 | 12 | 2.4% | 0.30 [-4.11, 4.71] |
| Kyung 2020 | 25.7 | 7.5 | 90 | 23.3 | 2.69 | 90 | 17.0% | 2.40 [0.75, 4.05] |
| Mapelli 2021 | 37.1 | 4.5 | 12 | 41.5 | 8 | 12 | 1.7% | -4.40 [-9.59, 0.79] |
| Roberge 2010 | 26.6 | 6.8 | 10 | 27.7 | 8.6 | 10 | 1.0% | -1.10 [-7.90, 5.70] |
| **Total (95% CI)** | | | **288** | | | **288** | **100.0%** | **1.72 [1.04, 2.40]** |

Heterogeneity: Chi² = 116.62, df = 8 (P < 0.00001); I² = 93%

Test for overall effect: Z = 4.97 (P < 0.00001)

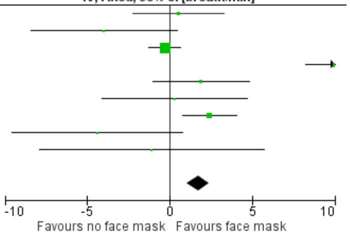


Supplementary Figure S8. Pooled analysis on the impact of surgical (A) and N95 (B) masks on respiratory rate.

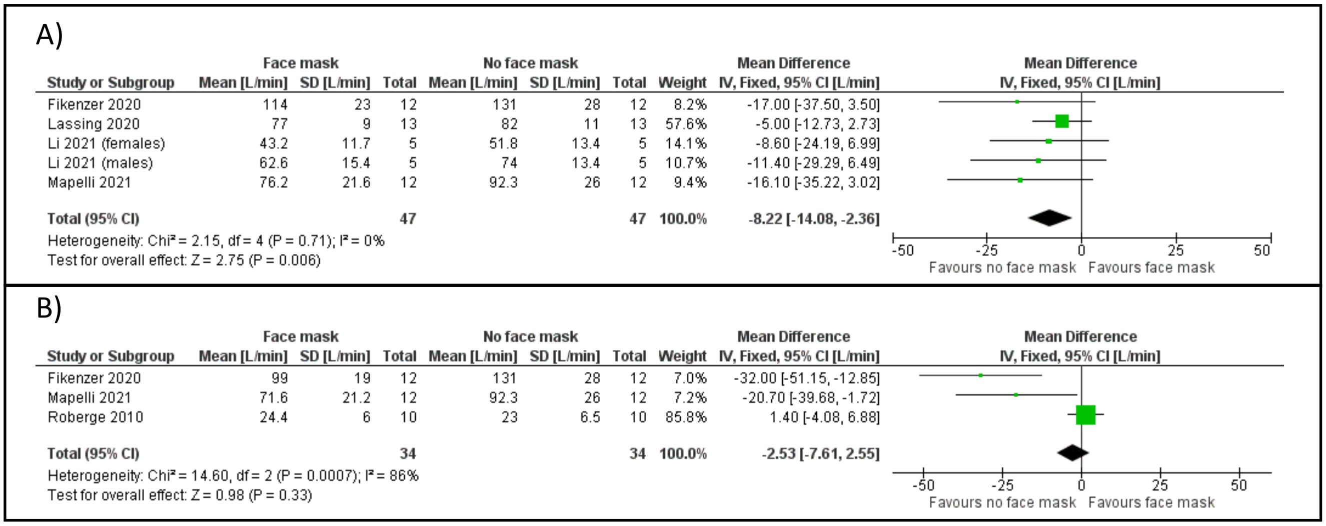


Supplementary Figure S9. Pooled analysis on the impact of surgical (A) and N95 (B) masks on ventilation.

A)

| Study or Subgroup | Face mask Mean [L] | Face mask SD [L] | Face mask Total | No face mask Mean [L] | No face mask SD [L] | No face mask Total | Weight | Mean Difference IV, Fixed, 95% CI [L] |
|---|---|---|---|---|---|---|---|---|
| Fikenzer 2020 | 2.9 | 0.5 | 12 | 3.2 | 0.7 | 12 | 13.3% | -0.30 [-0.79, 0.19] |
| Lassing 2020 | 2.5 | 0.4 | 13 | 2.5 | 0.4 | 13 | 33.3% | 0.00 [-0.31, 0.31] |
| Li 2021 (females) | 1.21 | 0.28 | 5 | 1.33 | 0.28 | 5 | 26.2% | -0.12 [-0.47, 0.23] |
| Li 2021 (males) | 1.9 | 0.4 | 5 | 2.23 | 0.31 | 5 | 16.0% | -0.33 [-0.77, 0.11] |
| Mapelli 2021 | 2.05 | 0.6 | 12 | 2.28 | 0.72 | 12 | 11.2% | -0.23 [-0.76, 0.30] |
| **Total (95% CI)** | | | **47** | | | **47** | **100.0%** | **-0.15 [-0.33, 0.03]** |

Heterogeneity: Chi² = 2.03, df = 4 (P = 0.73); I² = 0%
Test for overall effect: Z = 1.66 (P = 0.10)

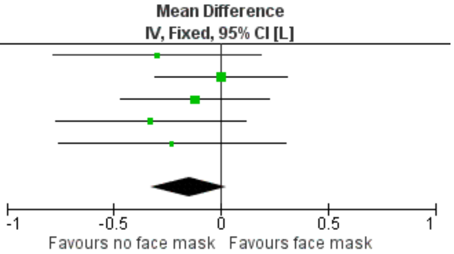


B)

| Study or Subgroup | Face mask Mean [L] | Face mask SD [L] | Face mask Total | No face mask Mean [L] | No face mask SD [L] | No face mask Total | Weight | Mean Difference IV, Fixed, 95% CI [L] |
|---|---|---|---|---|---|---|---|---|
| Fikenzer 2020 | 2.7 | 0.4 | 12 | 3.2 | 0.7 | 12 | 14.3% | -0.50 [-0.96, -0.04] |
| Mapelli 2021 | 1.96 | 0.65 | 12 | 2.28 | 0.72 | 12 | 9.8% | -0.32 [-0.87, 0.23] |
| Roberge 2010 | 0.95 | 0.24 | 10 | 0.86 | 0.21 | 10 | 75.9% | 0.09 [-0.11, 0.29] |
| **Total (95% CI)** | | | **34** | | | **34** | **100.0%** | **-0.03 [-0.21, 0.14]** |

Heterogeneity: Chi² = 6.56, df = 2 (P = 0.04); I² = 70%
Test for overall effect: Z = 0.39 (P = 0.70)

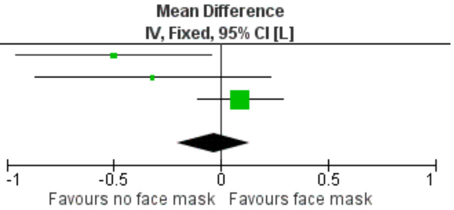


Supplementary Figure S10. Pooled analysis on the impact of surgical (A) and N95 (B) masks on tidal volume

A)

| Study or Subgroup | Face mask Mean [mmol/l] | Face mask SD [mmol/l] | Face mask Total | No face mask Mean [mmol/l] | No face mask SD [mmol/l] | No face mask Total | Weight | Mean Difference IV, Fixed, 95% CI [mmol/l] |
|---|---|---|---|---|---|---|---|---|
| Boldrini 2020 | 3.3 | 2.05 | 25 | 3.46 | 2.29 | 25 | 33.0% | -0.16 [-1.36, 1.04] |
| Fikenzer 2020 | 11 | 3.9 | 12 | 12.8 | 3.1 | 12 | 6.0% | -1.80 [-4.62, 1.02] |
| Lassing 2020 | 4.71 | 1.42 | 13 | 4.27 | 1.46 | 13 | 39.1% | 0.44 [-0.67, 1.55] |
| Ramos-Campo 2021 | 5.3 | 2.1 | 14 | 4.2 | 1.9 | 14 | 21.8% | 1.10 [-0.38, 2.58] |
| **Total (95% CI)** | | | **64** | | | **64** | **100.0%** | **0.25 [-0.44, 0.94]** |

Heterogeneity: Chi² = 3.85, df = 3 (P = 0.28); I² = 22%
Test for overall effect: Z = 0.71 (P = 0.48)

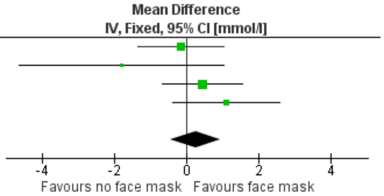


B)

| Study or Subgroup | Face mask Mean [mmol/l] | Face mask SD [mmol/l] | Face mask Total | No face mask Mean [mmol/l] | No face mask SD [mmol/l] | No face mask Total | Weight | Mean Difference IV, Fixed, 95% CI [mmol/l] |
|---|---|---|---|---|---|---|---|---|
| Fikenzer 2020 | 10.8 | 3.1 | 12 | 12.8 | 3.1 | 12 | 30.5% | -2.00 [-4.48, 0.48] |
| Ramos-Campo 2021 | 4.5 | 2.5 | 14 | 4.2 | 1.9 | 14 | 69.5% | 0.30 [-1.34, 1.94] |
| **Total (95% CI)** | | | **26** | | | **26** | **100.0%** | **-0.40 [-1.77, 0.97]** |

Heterogeneity: Chi² = 2.29, df = 1 (P = 0.13); I² = 56%
Test for overall effect: Z = 0.58 (P = 0.56)

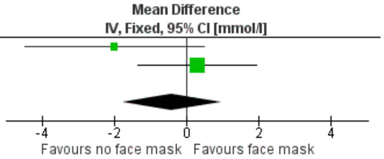


Supplementary Figure S11. Pooled analysis of the effects of surgical (A) and N95 (B) masks on lactate.
