## Supplementary Figure for "The Impact of Face Masks on Performance and Physiological Outcomes during Exercise: A Systematic Review and Meta-analysis"

**Supplementary Table S1.** Risk of bias in studies involving face masks during exercise

| Study |  | Risk of Bias Domain | | | | | |
| --- | --- | --- | --- | --- | --- | --- | --- |
|  | Randomization process | Period or carry-over effect | Deviation from intended intervention | Missing outcome data | Measurement of outcome | Selection of reported results | Overall risk of bias |
| Barbeito-Caamaño et al., 2021 | High | Some concerns | Low | Low | Low | Some concerns | High |
| Boldrini et al., 2020 | Some concerns | Some concerns | Low | Low | Low | Some concerns | Some concerns |
| Carrizal & Rodríguez, 2020 | High | Some concerns | Low | Low | Low | Some concerns | High |
| Epstein et al., 2020 | Some concerns | Some concerns | Low | Low | Low | Some concerns | Some concerns |
| Fikenzer et al., 2020 | Some concerns | Some concerns | Low | Low | Low | Some concerns | Some concerns |
| Goh et al., 2019 | High | High | Low | Low | Low | Low | High |
| Jones, 1991 | Some concerns | Low | Low | Low | Low | Some concerns | Some concerns |
| Kim et al., 2013 | High | High | Low | Low | Low | Some concerns | High |
| Kim et al., 2016 | Some concerns | Some concerns | Low | Low | Low | Some concerns | Some concerns |
| Kyung et al., 2020 | High | High | Low | Low | Low | Some concerns | High |
| Laird et al., 2002 | High | High | Low | Low | Low | Some concerns | High |
| Lässing et al., 2020 | Some concerns | Low | Low | Low | Low | Some concerns | Some concerns |
| Li et al., 2021 | Some concerns | Low | Low | High | High | Some concerns | High |
| Mapelli et al., 2021 | Low | Low | Low | Low | Low | Some concerns | Some concerns |
| Person et al., 2018 | Some concerns | Some concerns | Low | Low | Low | Some concerns | Some concerns |
| Ramos-Campo et al., 2021 | Some concerns | Low | Low | Low | Low | Some concerns | Some concerns |
| Roberge et al., 2010 | Some concerns | High | Low | Low | Low | Some concerns | High |
| Roberge et al., 2012a | High | High | Low | Low | Low | Some concerns | High |
| Roberge et al, 2012b | High | High | Low | Low | Low | Some concerns | High |
| Shaw et al., 2020 | Low | Low | Low | Low | Low | Low | Low |
| Shein et al., 2021 | High | High | Low | Low | Low | Some concerns | High |
| Wong et al., 2020 | Some concerns | High | Low | Low | Low | Some concerns | High |
